# Homelessness, type of homelessness, and risk of cause-specific mortality: a systematic review and meta-analysis of 116 studies comprising 2,563,633 homeless people and 129,292,553 population controls

**DOI:** 10.1101/2025.04.22.25326193

**Authors:** J White, Y Moriarty, M Lau, R Cannings-John, A Palmer, A L Weightman, M Kiseleva, G D Batty

**Affiliations:** Centre for Trials Research, School of Medicine, Cardiff University, Cardiff, UK; Centre for the Development, Evaluation, Complexity and Implementation in Public Health ImpRovement (DECIPHer), School of Social Sciences, Cardiff University, Cardiff, UK; Specialist Unit for Review Evidence (SURE), Cardiff University, Cardiff, UK; Department of Epidemiology and Public Health, University College London, London, UK

## Abstract

**Background:** Homelessness might increase the risk of premature mortality, but evidence is scarce, imprecise, and is mostly limited to rough sleepers as opposed to more common types of homelessness.

**Methods:** Published studies were retrieved through a systematic search of MEDLINE, Embase PsycINFO and Scopus from inception to December 2024. Unpublished data were identified from open-access data archives. We used random-effects meta-analysis to combine effect estimates from published and unpublished data. This review is registered at PROSPERO (CRD42023430984).

**Findings:** We included 116 studies from Europe, the USA, South America, Africa, Asia, and Australia. The meta-analysis of all-cause mortality comprised 110,892,271 people (1,618,049 exposed to homelessness). The risk of all-cause mortality was significantly increased in people exposed to homelessness (Relative risk [RR] 2·12 [95% CI 1·91-2·57], p<0·001, I^2^=99·7%), with risks similar in men (3·88, 2·69-5·06) and women (3·46, 2·17-4·70). This risk was most elevated in people who had slept rough (7·63, 3·29-11·97), followed by those who used low-cost hotels (5·18, 1·14-9·23), then hostels (3·44, 2·10-4·77). In analyses of cause-specific mortality (26,291,900 people, 1,202,205 homeless), summary RR estimates were elevated for 33 of the 36 (92%) causes of death and highest for deaths due to psychoactive substance use disorder (21·36, 14·44-31·67), accidental injuries (13·15, 5·46-31·69), drug-overdose (10·80, 6·37-18·31), and those that are alcohol-related (5·93, 1·10-22·04). No evidence of publication bias was detected.

**Interpretation:** Homeless people experience an increased risk of premature mortality across an array of health outcomes. That the most extreme inequities have an interrelated aetiology suggests a cross-sectoral medical, housing, and social care response is required.

**Funding:** The Centre for Homelessness Impact, Health and Care Research Wales, UK Medical Research Council (MR/P023444/1) and the US National Institute on Aging.

**Research in context:** *Evidence before this study:* While existing systematic reviews have found that homelessness is associated with an increased risk of all-cause mortality, there remains substantial gaps in knowledge. Currently there is no meta-analysis of the risk of death from all-causes and specific disorders, comparison of mortality risk across the spectrum of homelessness experience (e.g., rough sleeping, hostels, single-occupancy low-cost hotels), whether differences exist in the impact on men and women, or in high, or low and middle income countries (LMICs). Accordingly, we searched MEDLINE, Embase, PsycINFO and Scopus from inception to December 2024 for studies reporting mortality outcomes among people with a history of homelessness. Additionally, we searched the grey literature and obtained unpublished individual-participant data for two cohort studies from open-access data archives.

*Added value of this study:* Our systematic review and meta-analysis provides the first comprehensive examination to date of mortality among people who have experienced homelessness. Drawing on 116 studies, there was, in aggregate, a doubling in risk of all-cause mortality in the 1,618,049 people who had experienced homelessness relative to the 109,274,222 population controls (relative risk [RR] 2·12, [95% CI 1·91-2·57], p<0·001, *I*^2^=99·7%). This association was strongest in people who had slept rough (7·63, 95% CI 3·29-11·97) than in a hostel (3·44, 95% CI 2·10-4·77) and was of equal magnitude in men (3·88, 95% CI 2·69-5·06) and women (3·43, 95% CI 2·10-4·70). Summary RRs were extremely high for some causes of death, including RRs of >20 for psychoactive substance use disorder, 10-19 for drug-overdose, accidents, and accidental injuries, and 5-10 for alcohol-related causes - none of which have been previously summarised. No evidence of publication bias was found.

*Implications of all the available evidence:* Homelessness is associated with an increased risk of all-cause mortality. Our analysis suggests these a large proportion of these excess deaths are caused by substance use, drug-overdose, accidents and harmful alcohol consumption. The extreme inequalities in mortality we identified points to the need a cross-sectoral response to improve both health and housing conditions for people who have experienced homelessness.

## Background

Worldwide, it is estimated that nearly 1.6 billion people have inadequate shelter.^1^ In high-income countries, insufficient social and unaffordable private housing are leading to a surge in homelessness. An estimated 895,000 people in Europe – one in every 600 people – sleep rough in shelters or other temporary accommodation every night; an increase of 40% since 2023-24.^2^ Corresponding data from the US suggest around 771,480 people are experiencing homelessness, with the majority living in emergency shelters or temporary accommodation; up 18% from 2023-24. ^3^ In England, local government spending on emergency housing for homeless families has almost doubled, from £411 million in 2022-23 to an estimated £732 million the following year.^4^

While existing systematic reviews have provided an important evidence base,^5, 6^ gaps in understanding and methodological limitations hamper their usefulness for policy makers. First, there is currently no meta-analysis. The one previous meta-analysis combined mortality-risk in three studies with people who had been homeless with the people who had been incarcerated, a sex worker, and had a substance use disorder.^5^ Second, mortality risk across the spectrum of homelessness experience, which includes rough sleeping, use of hostels, and low-cost temporary accommodation, is not known. Third, while homelessness is more common in men,^7^ whether sex differences exist in its impact on mortality is untested. Fourth, previous reviews did not include studies from low- and middle-income countries. Lastly, publication bias - the notion that studies reporting positive results are more likely to be published - has not been explored and has the capacity to distort the evidence base.

We did this meta-analysis of the association between homelessness with all-cause and cause-specific mortality to address these gaps.

## Methods

The review was prospectively registered with PROSPERO (CRD42023430984)^8^ and conducted in accordance with a published protocol.^9^ We followed the PRISMA-IPD^10^ and MOOSE ^11^ reporting guidelines in the presentation of our report.

### Search strategy and selection criteria

We identified published studies through a systematic search of MEDLINE, Embase, PsycINFO and Scopus from their inception to December 2024. Full search terms are provided in the appendix (pp. 2). No limits on language were applied and articles were translated into English if required.

After the exclusion of duplicate studies, MK and AW independently reviewed all titles and abstracts of the remaining articles to establish their eligibility on the basis of defined inclusion criteria.^9^ Articles had to report estimates of mortality in people who were currently or who had ever been exposed to homelessness. Full texts were then retrieved of papers that appeared to meet the inclusion criteria, or where a decision could not be made, based on titles and abstracts. We also conducted a supplementary search of reference sections of systematic reviews and websites to identify grey literature. We excluded studies where a residence was (or had been) threatened due to financial insecurity; if they resided in poor living standards; or if report authors provided insufficient information to calculate an estimate, such as those studies solely quantifying mortality in a homeless population without an unexposed comparator group. Any disagreements were resolved through discussion, involving a third reviewer where necessary.

We identified duplicate data presented in separate publications. Where this occurred, we selected the study with the largest or most representative sample size, and when these were also similar, we used the most recently published report.

### Data extraction

All authors contributed to data extraction and all extracted information was double-checked. Extracted characteristics included: name of the first author, publication year, study location (country), study type, homelessness type, current or historical homelessness, comparator population, degree of socioeconomic disadvantage, follow-up period, number of people at risk and number of deaths in exposed and non-exposed groups, the summary statistic reported for age and sex (e.g. either reported as frequency, percentage or mean or median alongside standard deviation or interquartile range), and extent of statistical adjustment.

### Unpublished individual-participant data

We supplemented data from the published studies with unpublished individual-level data from prospective cohort studies, as located by searching the UK Data Service (https://ukdataservice.ac.uk) and the Inter-University Consortium for Political and Social Research (http://www.icpsr.umich.edu/icpsrweb/ICPSR/). This yielded two criteria-meeting cohort studies: the Survey of Health, Ageing and Retirement in Europe^12^ and the US Health & Retirement Study.^13^ In both, data collection and access had been approved by the relevant local or national ethics committee and all participants gave informed consent to participate. Harmonised covariates included age, sex, and socioeconomic status.

### Outcomes

Outcomes were all-cause and cause-specific mortality defined according to the International Classification of Diseases, tenth revision (ICD-10).

### Quality assessment

To assess the quality of included studies, we used the MethodologicAl STandards for Epidemiological Research (MASTER) scale.^14^ Assessments were made across seven domains: formal recruitment, equal retention, equal ascertainment, equal implementation, equal prognosis, sufficient analysis and temporal precedence (minimum score = 0, maximum (high quality) = 36). The quality of the study was regarded as adequate (i.e., low risk of bias) if the total score was above a median of 12.

### Statistical analysis

The statistical analysis plan was completed before analysis (appendix 3, pp 6). The data and code for all the statistical analyses can be found on GitHub: https://github.com/jameswhite1979/Homelessness-systematic-review. We first analysed unpublished individual-participant data using Cox proportional hazards regression to generate hazard ratios (HRs) with accompanying 95% confidence intervals (CIs) for the associations between homelessness, all-cause, and cause-specific mortality.^15^

Next, we used meta-analysis to combine the results from the analyses of the unpublished data with estimates from published studies. Where published studies provided HRs and odds ratios (ORs), these were used as a common estimate of relative risk (RR). In studies where the incidence of mortality was <10%, we regarded ORs as a close approximations of RR.^16^ If mortality incidence was ≥10% and count data were available, then ORs and HRs were converted to RRs.^17,18^ If confidence intervals were missing, they were calculated using standard formulae.

We analysed the associations of homelessness with all-cause and cause-specific mortality separately. For the unpublished data, effect estimates were adjusted for age and sex, and socioeconomic status. For published studies, we used the most comprehensively adjusted estimates. Some publications reported multiple effect sizes based on different comparator groups, so creating a unit-of-analysis issue. To account for this, when multiple comparator populations groups were reported, we used estimates where the comparison group was a socially deprived population, or where measures were adjusted for area, household, or an individual-level deprivation. If studies reported multiple comparisons (e.g., for short and longer periods of follow-up) we chose the comparison with the greatest number of deaths. Where multiple exposures were compared to the same comparator group (e.g., different types of homelessness to the same unexposed population), exposures were combined using count data. Where the exposed group was compared to more than one comparator group (e.g., to another cohort participants or an external population) we ran a multilevel meta regression to model the effects nested within studies to account for the association between the common groups.^19^

We used random effects models reflecting the anticipated heterogeneity in the included studies. We used the *I^2^* statistic to summarise the proportion of total variation in study estimates due to heterogeneity, with higher values denoting greater heterogeneity.^20^ Planned subgroup analyses of the homelessness-mortality association were conducted in different populations including by participant sex, country, homelessness status (current *vs* historical), type of homeless (sleeping rough *vs* shelter *vs* temporary accommodation), and according to income of the country from which the study sample was drawn based on the World Bank 2023 classification.^21^ Subgroup analyses also examined differences according to the study characteristics of the method of exposure ascertainment (self-report *vs* administrative record), comparator population (socioeconomically deprived *vs* general; internal *vs* external cohort), effect sizes adjustment (minimal *vs* multivariable), publication status (unpublished *vs* published), study design (retrospective *vs* prospective), total sample size (less than 1k, 1k to <10k, 10k to <100k, 100k to <500k, 500k to <1m, 1m to <5m, 5m to <10m, 10m and over), and study quality (categorised low, medium, high). Subgroup differences were estimated using meta-regression.

Summary RRs for causes of death were first estimated at the ICD chapter level (e.g., cancer, external) and then higher specific cause resolution (e.g., lung cancer, suicide). Publication bias was explored using an analysis of asymmetry in funnel plots.^22^ Where detected, we used the non-parametric trim-and-fill method to quantify its effect.^23^ As we anticipated retrieving a small number of large studies, we used the ‘leave-one-out’ method^24^ to examine the impact of excluding each on the summary effect size.

We did statistical analyses with Stata (MP version 18) to analyse the unpublished data, and Stata (MP version 18) and R (version 4.3.3) to compute the meta-analysis.

## Role of the funding source

The funders of the study had no role in study design, data collection, data analysis, data interpretation, writing of the report, or the decision to submit the paper for publication. All authors had full access to all the data and had final responsibility for the decision to submit for publication.

## Results

Figure 1 depicts the process of study selection. We identified 7,630 records from four electronic database searches, of which 3,274 were duplicated. In a parallel supplementary search for eligible unpublished datasets and grey literature, we scrutinised the reference sections of eight systematic reviews, 12 websites and 16 online databases. In so doing, 36 additional records were identified. We reviewed the full texts of 350 studies, of which 116 (114 published plus two unpublished datasets) met our inclusion criteria, comprising at least 131**·**9 million people (appendix, pp. 5). Excluded studies and reasons for exclusion with study counts are listed in the appendix (pp. 9).

**Figure 1:**
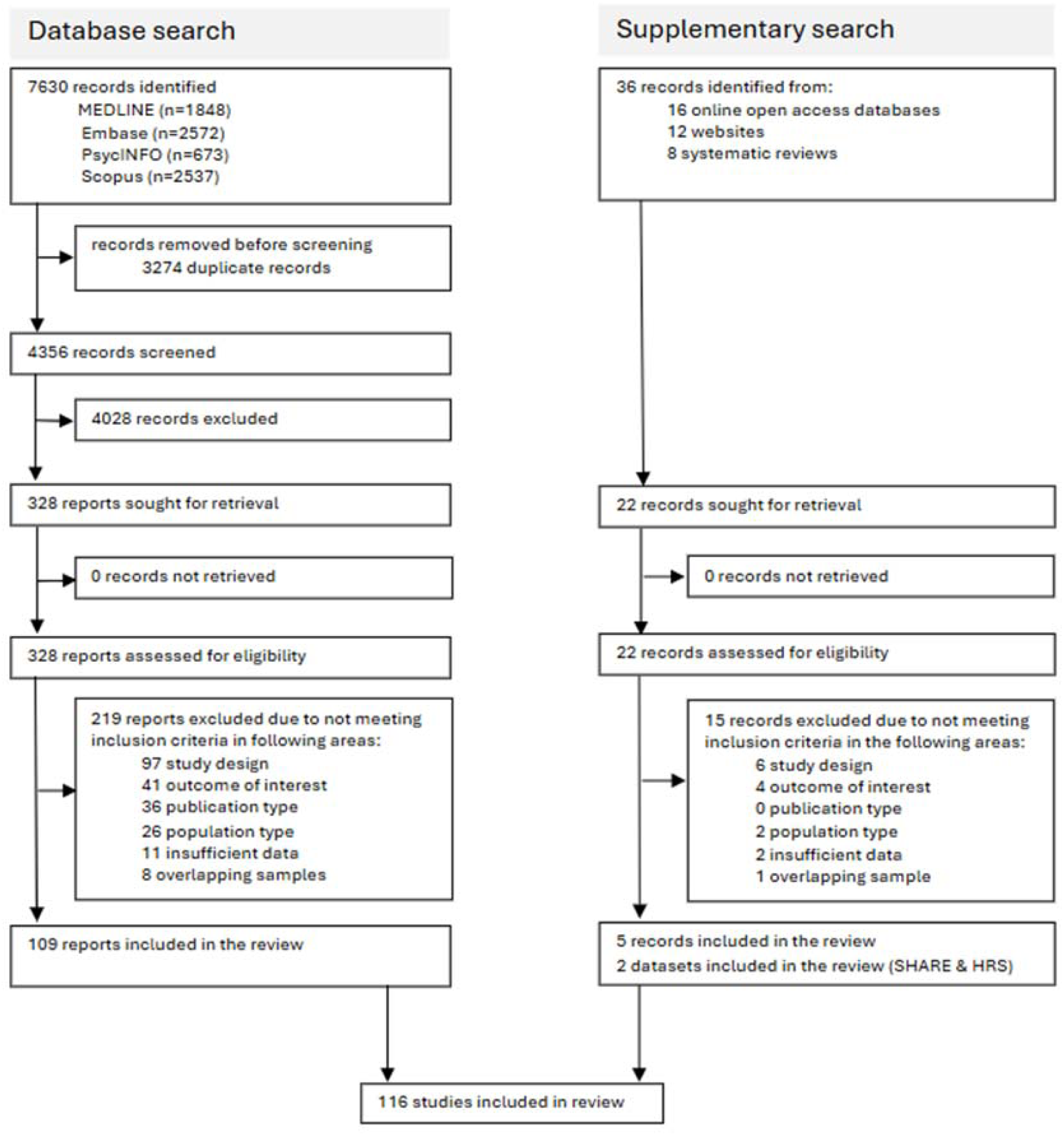
Study selection.

Included studies drew on samples from 22 countries: 55 studies from the USA; 32 from Europe (9 United Kingdom, 6 Netherlands, 3 Sweden, 4 Denmark, one each from Belgium, Finland, France, Greece, Ireland, Italy, Poland, Romania, and Spain, and one that included 13 European countries); 3 from Australia; 14 from Canada; 4 from South America; 3 from India; 2 from Russia; and 1 each from Kenya and Korea. Additionally, one study featured a combined sample from Estonia, Latvia, Philippines, Russia, and Peru. One hundred and twelve (96·5%) were from high income countries and four from LMICs. In the appendix (pp. 12), we provide study-level characteristics as well as the quality assessment of each. Of the 116 studies, 52 (44·8%) were evaluated as being of high quality. External causes of death were the most examined ICD-10 chapter-level cause, accounting for 65 (20·3%) of 320 computed effect sizes (appendix pp. 12).

A total of 1,618,049 people who were homeless and an estimated 109,274,222 from comparator populations contributed to the meta-analysis of all-cause mortality. Ninety-five studies yielded 126 unique RRs in the analyses of all-cause mortality, of which the large majority (91%, N= 115) had an elevated risk of death in the homeless group. Homelessness was associated with a doubling in the risk of all-cause mortality (RR 2·12, 95% CI 1·91–2·57, p<0·001; figure 1, appendix). There was, however, significant heterogeneity across studies with RR ranging between 0·17 and 37·3 (*I*^2^=99·7%; p<0·001, appendix). We found no strong evidence of publication bias (Egger’s test value = 1·86, p=0·06; figure 2, web appendix, pp.21).

**Figure 2:**
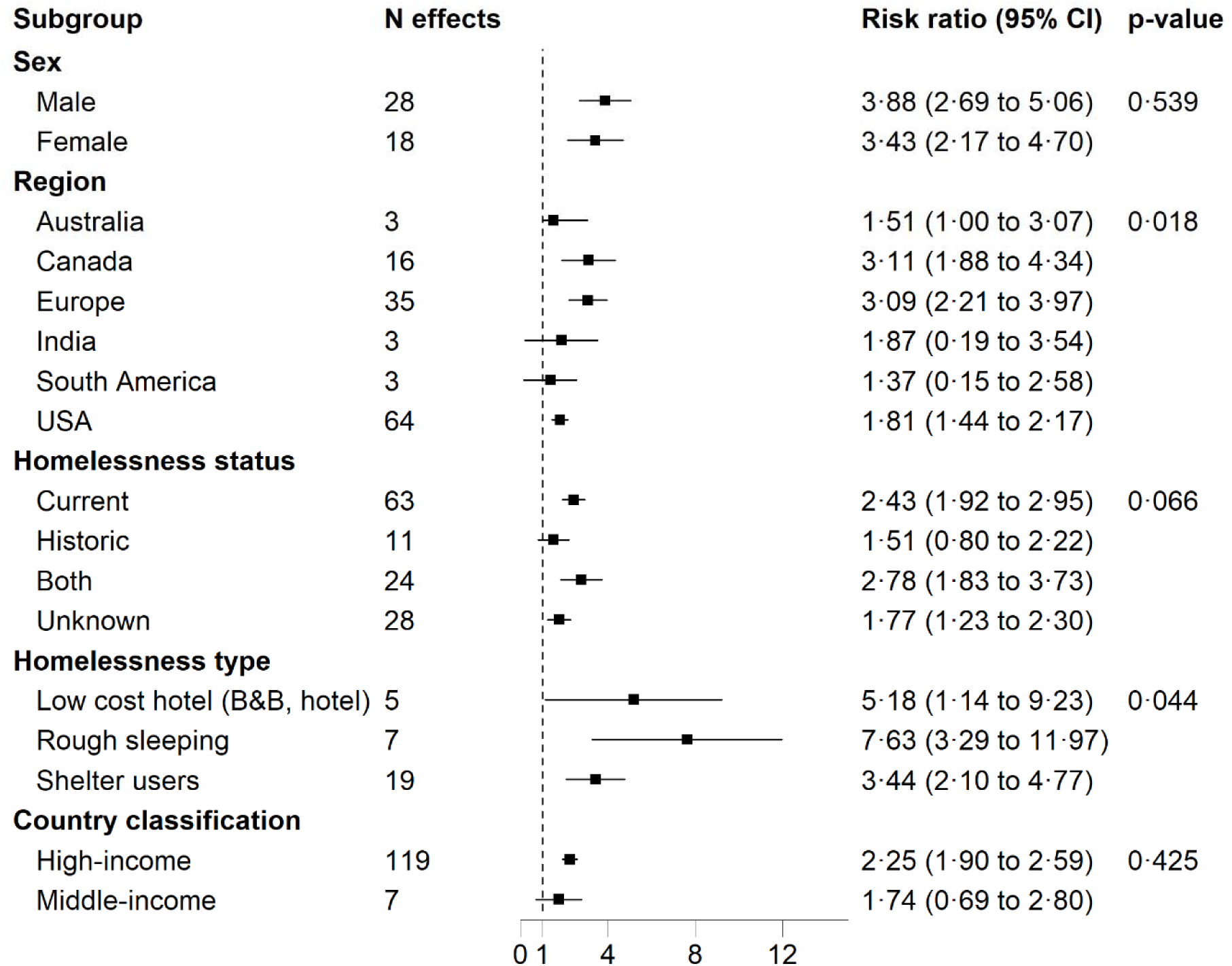
Association between homelessness and all-cause mortality comparing population subgroups. Estimates for unpublished data adjusted for age, sex, and socioeconomic status. For published studies, the most comprehensively adjusted estimates were included. Weights were assigned by random effects analysis. RR= relative risk.

Next, we examined the homelessness-mortality association across different population subgroups. There were no marked differences in this relationship between men (RR 3·88, 95% CI 2·69-5·06) and women (RR 3·43, 95% CI 2·17-4·70), or high (RR 2·25, 95% CI 1·90-2·59) compared to low income countries (RR 1·74, 95% CI 0·69-2·88). A stronger association was, however, apparent in people who had slept rough (RR 7·63, 95% CI 3·29-11·97) compared to using a shelter (3·44, 95% CI 2·10-4·77, p for difference = 0·04). There were also differences in effect estimate by geographical location, with higher RRs in samples drawn from Canada and Europe relative to the USA (p for difference = 0·02, figure 2).

Scrutinising the methodological characteristics of the studies, there was no strong evidence of bias arising from the type of comparator population used (general *vs* socioeconomically deprived); whether homelessness was current or historical; the method of exposure ascertainment; whether age was adjusted for in the model or not; or the study design. Summary RR for all deaths combined were, however, higher in studies which used an external compared to an internal comparator population; provided adjusted compared to unadjusted estimates; in lower grade studies; in studies with larger sample sizes (p for trend for increasing sample size < 0·001); and published studies had larger effects than unpublished studies (figure 3).

**Figure 3:**
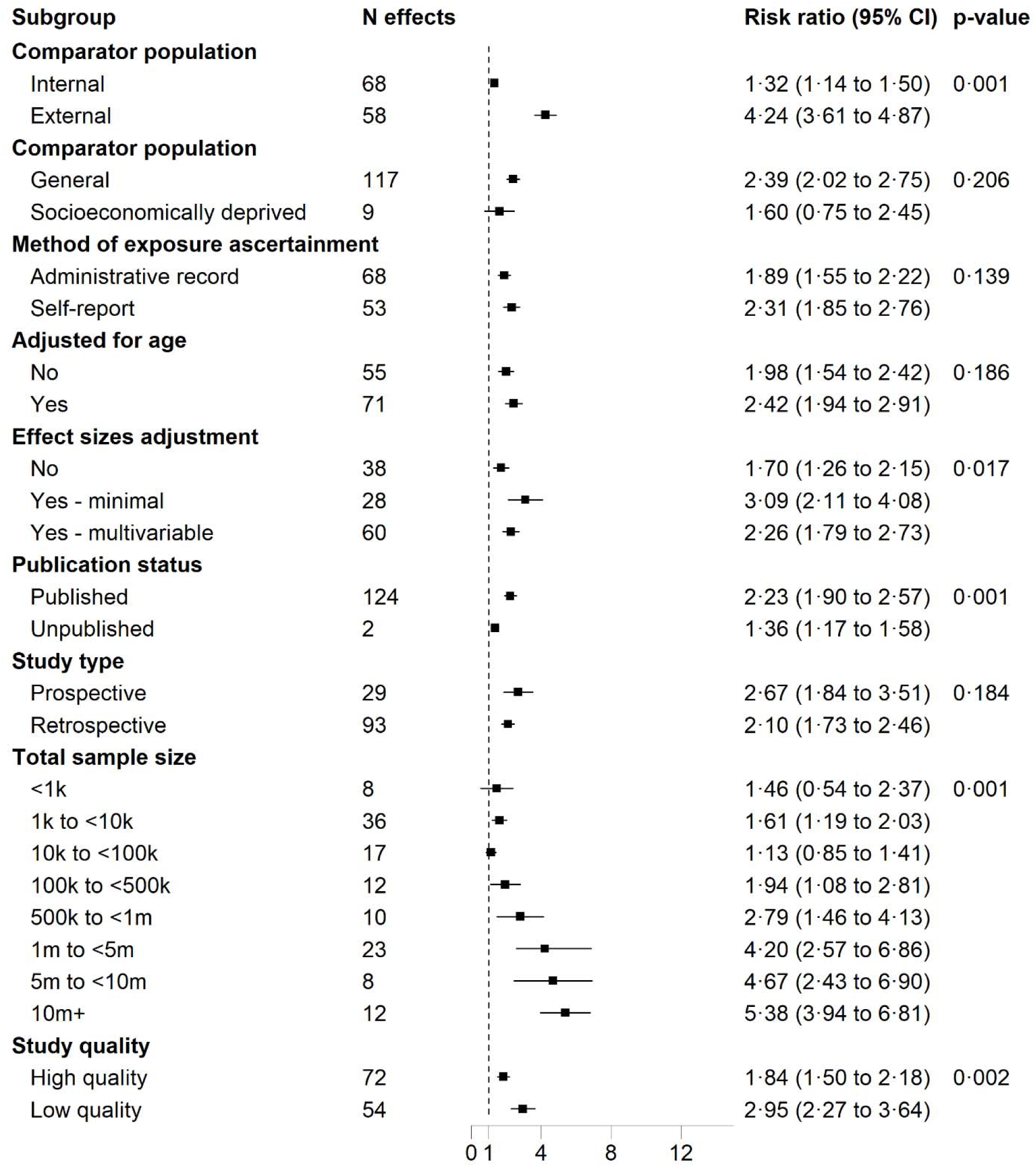
Association between homelessness and all-cause mortality comparing study characteristics. Estimates for unpublished data adjusted for age, sex, and socioeconomic status. For published studies, the most comprehensively adjusted estimates were included. Weights were assigned by random effects analysis. RR= relative risk.

The meta-analysis of homelessness and cause-specific mortality included 1,202,205 people who were homeless and an estimated 25,089,695 from comparator populations. Summary estimates for cause-specific mortality were elevated for 33 of the 36 (92%) causes of death investigated. In these analyses, summary RRs at the ICD-10 chapter level were particularly high for mental and behavioural disorders (8·51, 95% CI 4·99-14·51, F00 - F99 in ICD-10); injury, poisoning and other consequences of external causes (6·06, 95% CI 4·16-8·81, S00 -T98 in ICD-10); other/ unknown deaths (5·47, 95% CI 2·76-10·84, R00 - R99 in ICD-10); diseases of the blood and blood forming organs (5·37, 95% CI 1·19-24·17, D50 - D89 in ICD-10); and external causes (4·85, 95% CI 3·68-6·40, V01–Y98 in ICD-10) (figure 4).

**Figure 4:**
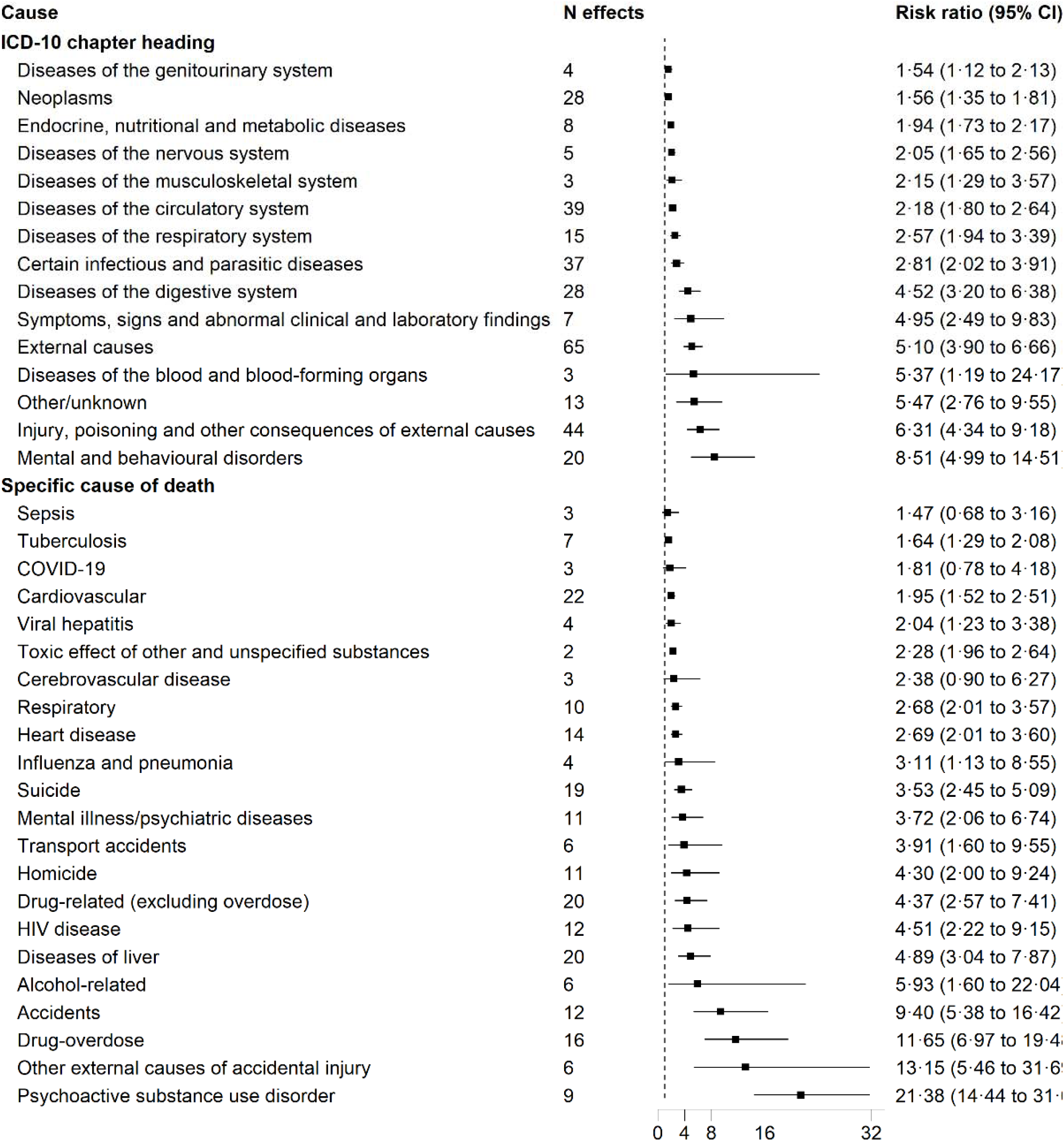
Association between homelessness and cause-specific mortality according to the ICD-10 disease categories. ICD-10 = International Classification of Diseases, tenth revision.

We then disaggregated cause of death from the ICD chapter level to individual causes. Summary RRs were highest for deaths attributed to psychoactive substance use disorder 21·36 (95% CI, 14·44-31·67); drug-overdose 10·80 (95% CI 6·37-18·31); other external causes of accidental injury 13·15 (95% CI 5·46-31·69); accidents 9·40 (95% CI 5·38-16·42); alcohol-related causes 5·93 (95% CI 1·60-22·04); symptoms, signs and abnormal clinical and laboratory findings, not elsewhere classified 4·95 (95% CI 2·49-9·83); drug-related (excluding overdose) 4·37 (95% CI 2·57-7·41); homicide 3·72 (95% CI 1·68-8·24); and suicide 3·35 (95% CI 2·30-4·87) (figure 4). Sensitivity analyses using the leaving-one-study-out approach to explore the effect of individual studies with larger sample sizes had little impact on these findings.

## Discussion

Our systematic review with meta-analysis of published and unpublished data revealed a series of new findings. We found a doubling in the risk of death in people with a history of homelessness relative to the general population, with the magnitude comparable for men and women, low and middle compared to high income countries. People who were exposed to the most severe end of the homelessness continuum – those sleeping on the street – were seven times and those in low cost hotels five times more likely to die than the general population. This mortality burden extended to almost all health conditions examined, with extreme RRs of >20 for psychoactive substance use disorder, 10-19 for drug-overdose and external causes of accidental injury, 5-10 for accidents and alcohol-related causes.

Our findings are consistent with the previous systematic reviews of homelessness and all-cause mortality^5,6^ both of which included a substantially smaller evidence base (11 studies) than in the present meta-analysis where 116 studies comprising at least 131**·**9 million people met our inclusion criteria. Importantly, for the first time to our knowledge, we provide a statistical aggregation of the association between homelessness across 36 causes of death. The available evidence was largest for external causes of death, but at the ICD-10 chapter level the risks of death were higher for mental and behavioural disorders, injury, and poisonings, and for specific causes were of a magnitude rarely seen in modern epidemiology to the extent that they support a causal link. We did not find any evidence of publication bias.

Our study has its strengths, including being the first meta-analysis of the association between homelessness and all-cause and cause specific mortality; its comparison of mortality risk across the spectrum of homeless experience and other population subgroups; and the inclusion of studies conducted in LMICs. Out work is not of course without its limitations. Some caution must be taken when interpreting the summary estimates because of the high degree of heterogeneity found between studies. The absence of agreed definitions of homelessness is likely to explain some of this variation. Some of the heterogeneity was associated with other methodological differences, with a stronger association apparent for all-cause in studies using external comparators, low study quality, and lacking control for potential confounders. Those studies that were had a limited availability of confounding factors were typically those generated from administrative records where often only age and sex were available. To further reduce the risk of confounding, we selected the most comprehensively adjusted estimate reported, and if multiple comparator populations were used, we selected the socioeconomically disadvantaged. Lastly, exposure to homeless, a time-varying exposure, was measured only once in the studies retrieved. Because people are likely to transition in and out of stable housing,^25^ our results might be subject to some misclassification whereby the captured studies have underestimated the true magnitude of the association.

Many of the individual causes of death we identified in people exposed to homelessness – psychoactive substance use disorder, drug overdose and those that are alcohol related – are eminently avoidable.^26^ Similarly, drug and alcohol misuse are likely to be implicated in other causes of mortality we found such as accidents and accidental injuries. This would suggest access to evidence-based treatments for substance use like methadone,^27^ standard or long acting buprenorphine maintenance therapy,^27^ the distribution of naloxone to treat opiate overdose, ^30^ and psychosocial interventions,^31^ may help ameliorate some of these excess deaths. Indeed, integration of these measures into rapid-response housing schemes for homeless people have been associated in short term improvements in hospital attendances, but not mental health, quality of life and substance use,^32^ suggesting additional approaches are required. Calls for an intersectoral response to inclusion health,^33^ where health, housing and social services work together to advocate for and deliver comprehensive services for people who are homeless should be empirically explored. In parallel, the lack of social housing and affordable private accommodation^34^ is a structural barrier that urgently needs to be addressed to enable policy responses to house people and support them to remain housed.

In conclusion, our meta-analysis shows that homeless is associated with around a doubling of the risk of all-cause mortality, with the most extreme inequalities found for deaths due to psychoactive substance use disorder, drug-overdose, accidental injury, and alcohol-related causes. The evidence in this review regarding the causes of death suggests more investment and evaluation of inclusion health policy responses integrating health, housing and social services is needed.

## Supporting information

Web appendix

## Data Availability

Individual participant data from the SHARE and HRS cohort studies are available to download from the Inter-University Consortium for Political and Social Research (http://www.icpsr.umich.edu/icpsrweb/ICPSR/). The data and code for the meta-analyses can be found on GitHub: https://github.com/jameswhite1979/Homelessness-systematic-review

https://github.com/jameswhite1979/Homelessness-systematic-review

## Contributors

JW & GDB generated the idea for this systematic review with all authors contributing to its development and the analytical plan. AW, MK, YM, and AP carried out the literature search and reviewed studies for inclusion. All authors extracted and checked the data. RCJ and ML conducted all the meta-analysis and JW wrote the first draft of the manuscript on which all authors contributed. All authors had full access to all the data in the study and had final responsibility for the decision to submit for publication.

## Declaration of interests

We declare no competing interests.

## Acknowledgments

We thank the study members from the SHARE and HRS cohort studies for their participation. This study was supported by funding from the Centre for Homelessness Impact (https://www.homelessnessimpact.org/). GDB is supported by the UK Medical Research Council (MR/P023444/1) and the US National Institute on Aging (1R56AG052519-01, 1R01AG052519-01A1), and JW by the Centre for Trials Research and The Centre for Development, Evaluation, Complexity and Implementation in Public Health Improvement (DECIPHer), both of which are supported by the Welsh Government through Health and Care Research Wales.

## Notes

### Competing Interest Statement

The authors have declared no competing interest.

### Clinical Protocols

https://osf.io/qtn7w/

### Author Declarations

Ethics review boards provided consent where relevant for all the studies included

