## Supplementary material for "Homelessness, type of homelessness, and risk of cause-specific mortality: a systematic review and meta-analysis of 116 studies comprising 2,563,633 homeless people and 129,292,553 population controls": Web appendix

**Web appendices**

### **Web appendix 1: Search terms**

**Ovid MEDLINE(R) ALL**

| 1 | homeless persons/ or homeless youth/ |
| --- | --- |
| 2 | ill-housed persons/ or emergency shelter/ |
| 3 | (homeless* or houseless* or unhoused).tw. |
| 4 | ((housing adj2 exclu*) or (housing adj2 cris*) or (housing adj2 concern*) or (housing adj2 instability) or (housing adj2 insecurity) or "residential instability" or "residential insecurity" or "no permanent address" or squat* or hostel* or sofa-surf* or couch-surf*).tw. |
| 5 | evict*.tw. |
| 6 | ((rough adj1 sleep*) or (sleep* adj3 street) or (slept adj3 street) or (liv* adj3 street) or "without a roof" or roofless).tw. |
| 7 | ((insecure* or unstable or non-permanent* or temporar* or improvised or emergency or marginal* or precarious* or nighttime or overnight or transitional* or recovery or crisis) adj (hous* or home* or accommodat* or dwell* or shelter* or residen*)).tw. |
| 8 | ((women* or youth or "young people*" or "young person*") adj shelter*).tw. |
| 9 | mortality/ or fatal outcome/ or survival rate/ or death/ |
| 10 | (mortalit* or death* or dying or fatal* or decease*).tw. |
| 11 | 1 or 2 or 3 or 4 or 5 or 6 or 7 or 8 |
| 12 | 9 or 10 |
| 13 | 11 and 12 |
| 14 | limit 13 to (comment or editorial or letter) |
| 15 | 13 not 14 |
| 16 | (interview* or "focus group*" or ethnograph* or single-case or "case report*" or "case stud*" or cross-sectional or case-control).tw. |
| 17 | 15 not 16 |

**Embase via Ovid**

| 1 | homeless person/ or homeless man/ or homeless woman/ or homeless youth/ or homelessness/ |
| --- | --- |
| 2 | housing instability/ |
| 3 | (homeless* or houseless* or unhoused).tw. |
| 4 | ((housing adj2 exclu*) or (housing adj2 cris*) or (housing adj2 concern*) or (housing adj2 instability) or (housing adj2 insecurity) or "residential instability" or "residential insecurity" or "no permanent address" or squat* or hostel* or sofa-surf* or couch-surf*).tw. |
| 5 | evict*.tw. |
| 6 | ((rough adj1 sleep*) or (sleep* adj3 street) or (slept adj3 street) or (liv* adj3 street) or "without a roof" or roofless).tw. |
| 7 | ((insecure* or unstable or non-permanent* or temporar* or improvised or emergency or marginal* or precarious* or nighttime or overnight or transitional* or recovery or crisis) adj (hous* or home* or accommodat* or dwell* or shelter* or residen*)).tw. |
| 8 | ((women* or youth or "young people*" or "young person*") adj shelter*).tw. |
| 9 | death/ or dying/ or fatality/ or mortality/ or all cause mortality/ or mortality rate/ |
| 10 | (mortalit* or death* or dying or fatal* or decease*).tw. |
| 11 | 1 or 2 or 3 or 4 or 5 or 6 or 7 or 8 |
| 12 | 9 or 10 |
| 13 | 11 and 12 |
| 14 | limit 13 to (editorial or letter) |
| 15 | 13 not 14 |
| 16 | (interview* or "focus group*" or ethnograph* or single-case or "case report*" or "case stud*" or cross-sectional or case-control).tw. |
| 17 | 15 not 16 |

**APA PsycInfo via Ovid**

| 1 | homeless/ or homeless mentally ill/ |
| --- | --- |
| 2 | shelters/ |
| 3 | (homeless* or houseless* or unhoused).tw. |
| 4 | ((housing adj2 exclu*) or (housing adj2 cris*) or (housing adj2 concern*) or (housing adj2 instability) or (housing adj2 insecurity) or "residential instability" or "residential insecurity" or "no permanent address" or squat* or hostel* or sofa-surf* or couch-surf*).tw. |
| 5 | evict*.tw. |
| 6 | ((rough adj1 sleep*) or (sleep* adj3 street) or (slept adj3 street) or (liv* adj3 street) or "without a roof" or roofless).tw. |
| 7 | ((insecure* or unstable or non-permanent* or temporar* or improvised or emergency or marginal* or precarious* or nighttime or overnight or transitional* or recovery or crisis) adj (hous* or home* or accommodat* or dwell* or shelter* or residen*)).tw. |
| 8 | ((women* or youth or "young people*" or "young person*") adj shelter*).tw. |
| 9 | "death and dying"/ or mortality rate/ or mortality risk/ |
| 10 | (mortalit* or death* or dying or fatal* or decease*).tw. |
| 11 | 1 or 2 or 3 or 4 or 5 or 6 or 7 or 8 |
| 12 | 9 or 10 |
| 13 | 11 and 12 |
| 14 | limit 13 to (comment or editorial or letter) |
| 15 | 13 not 14 |
| 16 | (interview* or "focus group*" or ethnograph* or single-case or "case report*" or "case stud*" or cross-sectional or case-control).tw. |
| 17 | 15 not 16 |

**Scopus**

TITLE-ABS ( ( ( homeless*  OR  houseless*  OR  unhoused  OR  ( housing  W/1  exclu* )  OR  ( housing  W/1  cris* )  OR  ( housing  W/1  concern* )  OR  ( housing  W/1  instability )  OR  (housing  W/1  insecurity )  OR  "residential instability"  OR  "residential insecurity"  OR  "no permanent address"  OR  squat*  OR  hostel*  OR  sofa-surf*  OR  couch-surf*  OR  evict*  OR  ( rough  W/0  sleep* )  OR  ( sleep*  W/2  street )  OR  ( slept  W/2  street )  OR  ( liv*  W/2  street )  OR  "without a roof"  OR  roofless  OR  ( (insecure*  OR  unstable  OR  non-permanent*  OR  temporar*  OR  improvised  OR  emergency  OR  marginal*  OR  precarious*  OR  nighttime  OR  overnight  OR  transitional*  OR  recovery  OR  crisis )  PRE/1  ( hous*  OR  home*  OR  accommodat*  OR  dwell*  OR  shelter*  OR  residen* ) )  OR  ( ( women*  OR  youth  OR  "young people*"  OR  "young person*" )  PRE/1  ( shelter* ) ) )  AND  ( mortalit*  OR  death*  OR  dying  OR  fatal*  OR  decease* ) )  AND NOT  (interview*  OR  "focus group*"  OR  ethnograph*  OR  single-case  OR  "case report*"  OR  "case stud*"  OR  "cross-sectional"  OR  "case-control" ) )  AND  ( EXCLUDE ( DOCTYPE ,  "no" )  OR  EXCLUDE ( DOCTYPE ,  "ed" )  OR  EXCLUDE ( DOCTYPE ,  "le" ) )

### **Web appendix 2: Description of the cohort studies with unpublished individual-participant data**

*Survey of Health, Ageing and Retirement in Europe*^1^

The Survey of Health, Aging and Retirement in Europe (SHARE) is an international European database containing detailed information about the demographics, health, and social and economic status from representative samples of community-based populations aged >50 years. The SHARE target population consists of all persons aged 50 years and over at the time of sampling who have their regular domicile in the respective SHARE country in Europe. A person is excluded if she or he is incarcerated, hospitalized or out of the country during the entire survey period, unable to speak the country’s language(s) or has moved to an unknown address. Of these, 24,276 men and women had data on exposure to homelessness and mortality and were eligible for our meta-analyses.

*Health & Retirement Study* ^2^

The Health and Retirement Study (HRS) is a nationally representative longitudinal survey of over 37,000 individuals aged 50 and older across 23,000 households in the USA. Conducted every two years since 1992, the study was designed as a national resource to track changes in health and economic conditions related to aging at both individual and population levels. It covers four key areas: income and wealth; health, cognition, and healthcare utilization; work and retirement; and family relationships. Of these, 7,250 men and women had data on exposure to homelessness and mortality and were eligible for our meta-analyses.

### **Web appendix 3: Statistical analysis plan**

In the main analyses of the unpublished individual-participant (raw) data, we used Cox proportional hazards models to generate hazard ratios (HRs) with accompanying 95% confidence intervals (CIs) as a summary of the association between homelessness and all-cause mortality in each of the available studies. Using meta-analytical procedures, we combined the results from these analyses of the unpublished data with the estimates from the published studies reported as HRs and odds ratio (OR) to produce a common estimate of relative risk (RR). Where mortality incidence was low in the cohort studies (<10%), we regarded unadjusted and adjusted ORs as close approximations of RRs.^3^ Where mortality incidence is ≥10% and counts available, then ORs were converted using VanderWeele’s method (using a square-root transformation).^4^ Similarly, HRs were converted another VanderWeele’s method.^5^ Where missing, confidence intervals will be calculated using standard formulae (1.96 x standard error of standardised mortality ratio (SMR) where the standard error of SMR = (square root of observed deaths) / number of expected deaths).

Where multiple-adjusted estimates are available, we used those from the most complex multivariable model. The basic model included either age, sex, or a measure of socioeconomic disadvantage covariates as a minimum. A fixed value of 0.5 was added to cells with zero events so that an estimate can be derived. ^6^

Several studies reported more than one effect size; for instance, where different types of homelessness are compared to the same comparator group or where the same exposure group was compared to more than one comparator group, and so on). This creates a unit-of-analysis error due to the unaddressed correlation between the estimated effects of homelessness. To avoid this bias in the meta-analysis, we followed a reductionist (resolving within studies, thereby removing it from the meta‐analysis) and integrative (allowing within‐study effect size differences to be explored but introduce statistical issues with regard to the dependent effect sizes) approaches suggested by Cochrane^6^ and López-López JA et al ^7^:

- Select one effect size and exclude others: Where studies reported nested outcomes (e.g. mortality reported over short and long time periods, both sexes combined and males and females separately), we chose the comparison with the greatest number of deaths for the main comparison.
- Combine groups to create a single pair-wise comparison: In the situation where several types of exposure were analysed (e.g. rough sleeping, sofa-surfing etc; homeless <1 year and 1 year or more), the overarching ‘ever homeless’ exposure was used for the main comparison. Where applicable, the individual effects were used in subgroup analyses (e.g. by type of homelessness, individual sex effects, socioeconomically disadvantaged, or internal vs external comparator groups).
- Where an exposed group is compared to more than one comparator group (e.g. compared to two general populations or compared to an internal and external population) or vice-versa, more than one exposure group will be compared to a comparator, we accounted for these association by running a multilevel meta regression including a random effects for these effects nested within studies.

An important distinction is between situations in which a study can contribute several *independent* comparisons (i.e. with no exposure or comparator group in common); it is usually reasonable to include independent comparisons in a meta-analysis as if they were from different studies. In studies where homeless vs non-homeless comparators were examined in different patient groups (e.g. older and younger age groups, different time periods) we treated effect estimates as independent for the purposes of the meta-analysis.

Forest plots were used to visually depict summarised differences between homeless individuals and comparator populations. Random effects models were used, reflecting the anticipated heterogeneity in the studies included of different studies and populations. We used the restricted maximum likelihood (REML) method to estimate the heterogeneity variance as recommended by the Cochrane statistical methods group.^8^ Heterogeneity was computed using the I² statistic and reported alongside all meta-analyses.^33^ Values of I² >50% denoted moderate and >75% substantial heterogeneity.^34^

Potential publication bias was examined using Egger’s regression-based test with a p-value < 0.05 indicating significant asymmetry and that bias may be a problem. When publication bias was detected, we used the non-parametric trim-and-fill method proposed by Duval & Tweedie to quantify the effect of this publication bias.^35^ We used the ‘leave-one-out’ meta-analysis, to examine the influence of impact of excluding one study at each analysis (influential studies), on the overall effect -size estimate.

We explored the magnitude of the homelessness–mortality association according to different contexts, including sex, country as well as whether they are a high, medium or low-income country, study design, estimates previously unpublished or published, grey vs peer reviewed literature, total sample size, complexity of covariate adjustment (age adjusted vs non-age adjusted effects, minimal and multivariable adjusted effect sizes), type of comparator population (socioeconomically disadvantaged or not; internal - within study or external with comparison group from the general population), current or historical experience of homelessness (or both, unknown), type of homeless experience, method of reporting homelessness (self-report, administrative record). We also examine the impact of study quality based on quality assessment scores from the MASTER scale^9^ from which we dichotomised at the median score to create a high vs medium/ low quality subgroup. Forest plots will again be used to summarise multiple sub-group analyses.

We analysed associations of homelessness with ICD-10 chapter-level and individual level causes.

We used Meta Set to declare the pre-computed effect sizes.^10^ In accordance with the Meta-Analysis of Observational Studies guidelines,^11^ we used all available data in the main analysis and carried out sensitivity analysis including only high-quality studies according to the assessment of bias. All analyses will be conducted in (MP version 18) and R (version 4.3.3)

The data and code for these analyses can be found on GitHub: <https://github.com/jameswhite1979/Homelessness-systematic-review>

### **Web appendix 4: Excluded studies at full text review**

| # | Author | Year | Source | Reason for exclusion |
| --- | --- | --- | --- | --- |
| 1  2  3  4  5  6  7  8  9  10  11  12  13  14  15  16  17  18  19  20  21  22  23  24  25  26  27  28  29  30  31  32  33  34  35  36  37  38  39  40  41  42  43  44  45  46  47  48  49  50  51  52  53  54  55  56  57  58  59  60  61  62  63  64  65  66  67  68  69  70  71  72  73  74  75  76  77  78  79  80  81  82  83  84  85  86  87  88  89  90  91  92  93  94  95  96  97  98  99  100  101  102  103  104  105  106  107  108  109  110  111  112  113  114  115  116  117  118  119  120  121  122  123  124  125  126  127  128  129  130  131  132  133  134  135  136  137  138  139  140  141  142  143  144  145  146  147  148  149  150  151  152  153  154  155  156  157  158  159  160  161  162  163  164  165  166  167  168  169  170  171  172  173  174  175  176  177  178  179  180  181  182  183  184  185  186  187  188  189  190  191  192  193  194  195  196  197  198  199  200  201  202  203  204  205  206  207  208  209  210  211  212  213  214  215  216  217  218  219  220  221  222  223  224  225  226  227  228  229  230  231  232  233  234 | Adams  Adams  Agarwal  Akambase  Aldridge  Alpert  Alstrom  Alstrom  Altamura  Andreev  Arghir  Arnautovska  As  Asgary  Auerswald  Austin  Babidge  Baez-Saldana  Baggett  Baggett  Baggett  Balasuriya  Balla  Barocas  Bauer  Benzano  Bickley  Bjorkenstam  Blosnich  Burman  Boivin  Booth  Borg  Bradford  Brakefield  Brown  Calvo  Cavanaugh  Cawley  Center for Disease Control  Center for Disease Control  Center for Disease Control  Cha  Chandra  Chang  Chant  Charvin-Fabre  Che  Chen  Chenciner  Chiu  Choi  Clark  Concannon  Corbet  Corey  Cucher  Culatto  Cuttler  Di Gennaro  Di Gennaro  Dickins  Dos Santos  Embleton  Ezeani  Fan  Fica  Fine  Flach  Fluharty  Fowle  Fowler  Fuller  Funk  Fuster  Gambatese  Genberg  Gershon  Gjersing  Glynn  Goverment gateway account  Guthmann  Hall  Hall  Hassanally  Hatamabadi  Helpman  Henkind  Hessol  Hewett  Hickox  Hoffberg  Holliday  Hood  Hoshino  Hsu  Huang  Huff  Husain  Hwang  Ivers  Jahiel  Jones  Jones  Joo  Kawatsu  Khan  Khan  Kibel  Knoble  Knowles  Kostadinov  Lachaud  Lane  Lebrun-Harris  Lee  Lee  Leifheit  Levin  Lewer  Lim  Lin  Liu  Lozada Ramos  Marks  Marshall  McCarthy  McLaughlin  McMahon  McMillan  McMillan  Metraux  Mitchell  Molina  Montgomery  Montgomery  Montgomery  Montgomery  Moore-Pardo  Morrison  NY City Health  NY City Dep. Health and Mental Hygiene  Nielsen  Nilsson  Nishimura  Nordentoft  Nordentoft  Nouri  Nusselder  Obialo  O’Connell  O’Connor  Ohsaka  Oliveira  Onyeka  Osman  Oud  Padwa  Page  Pagidpally  Park  Park  Park  Patel  Patricio  Paudyal  Peek-Asa  Powell  Quilty  Radunovi  Rahai  Roberts  Roberts  Rodriguez  Rojas  Romaszko  Roncarati  Roncarati  Roncarati  Routhier  Routhier  Routhier  Routhier  Rowe  Roy  Roy  Russell  Saenko  Salindri  Samorodskaya  Santos  Scheuer  Schinka  Schinka  Shanks  Silver  Simone  Sinyor  Sinyor  Skrahina  Slockers  Slockers  Smetanina  Smith  Song  Spittal  Stenius-Ayoade  Suzuki  Suzuki  Swamy  Termorshuizen  Thakarar  Thomas  Thomas  Thomas  Thornley  Tierney  Tinl  van Draanen  Villa  Vrouwe  Vuillermoz  Vuillermoz  Youssef  Wainman  Waugh  Wingert  Wlodarczyk  Wright  Wu  Yu  Zhao  Zygmut  Zygmut | 2007  2023  2018  2024  2018  1997  1974  1975  2002  2008  2018  2014  1980  2018  2016  2021  2001  2016  2011  2015  2015  2020  2020  2019  2016  2021  2006  2017  2020  1997  2005  2024  1976  2016  2022  2022  2021  2012  2022  1987  1987  1991  2021  2024  2023  2013  2020  2021  2022  2021  2019  2011  2013  2019  2023  2022  2024  2021  2009  2022  2022  2023  2021  2018  2024  2022  2019  2021  2022  2021  2024  2015  2021  2022  2014  2013  2019  2019  2018  2017  2021  2020  2012  2019  2018  2019  2022  2023  2019  2011  2014  2018  2022  2019  2006  2020  2023  2022  2021  2002  2019  1992  2013  2022  2017  2014  2020  2024  2020  2016  2019  2020  2017  2018  2013  2024  2024  2021  2021  2020  2015  2022  2020  2021  2011  2019  1966  2023  2011  2014  2015  2011  2022  2022  2016  2017  2021  2024  2019  2009  2005  2010  2011  2014  2019  2007  2004  2022  2014  2005  2008  2014  2003  2020  2016  2023  2018  2022  2012  2023  1996  2022  2024  2023  2016  2021  2021  2023  2016  2017  2021  2022  2023  2022  2017  2017  2020  2021  2021  2017  2018  2020  2021  2019  1998  2004  2021  2020  2018  2017  2023  2018  2016  2018  1984  2023  2022  2015  2017  2015  2015  2018  2022  2021  2022  2006  2018  2013  2017  2024  2014  2019  2011  2021  2023  2016  2022  2021  2020  2014  2020  2014  2016  2024  2012  2018  2021  1992  1995  2018  2015  2022  2020  2020 | Database searches  Database searches  Database searches  Database searches  Database searches  Database searches  Database searches  Database searches  Database searches  Database searches  Database searches  Database searches  Database searches  Database searches  Database searches  Database searches  Database searches  Database searches  Database searches  Database searches  Database searches  Database searches  Database searches  Database searches  Database searches  Database searches  Database searches  Database searches  Database searches  Database searches  Database searches  Database searches  Database searches  Database searches  Database searches  Database searches  Database searches  Database searches  Database searches  Database searches  Database searches  Database searches  Supplementary search  Database searches  Database searches  Database searches  Database searches  Database searches  Database searches  Database searches  Database searches  Database searches  Database searches  Database searches  Database searches  Database searches  Database searches  Database searches  Database searches  Database searches  Database searches  Database searches  Database searches  Database searches  Database searches  Database searches  Database searches  Database searches  Database searches  Database searches  Database searches  Database searches  Database searches  Database searches  Database searches  Database searches  Database searches  Database searches  Database searches  Database searches  Database searches  Database searches  Database searches  Database searches  Database searches  Database searches  Database searches  Database searches  Database searches  Database searches  Database searches  Database searches  Database searches  Database searches  Database searches  Database searches  Database searches  Database searches  Database searches  Database searches  Database searches  Database searches  Database searches  Database searches  Database searches  Database searches  Database searches  Database searches  Database searches  Database searches  Database searches  Database searches  Database searches  Database searches  Supplementary search  Database searches  Database searches  Database searches  Database searches  Database searches  Database searches  Database searches  Database searches  Database searches  Database searches  Database searches  Database searches  Database searches  Database searches  Database searches  Database searches  Database searches  Database searches  Database searches  Database searches  Database searches  Database searches  Database searches  Database searches  Database searches  Supplementary search  Supplementary search  Database searches  Database searches  Database searches  Database searches  Database searches  Database searches  Database searches  Database searches  Database searches  Database searches  Database searches  Database searches  Supplementary search  Database searches  Database searches  Database searches  Database searches  Database searches  Database searches  Database searches  Database searches  Database searches  Database searches  Database searches  Database searches  Database searches  Database searches  Database searches  Database searches  Database searches  Database searches  Database searches  Database searches  Database searches  Database searches  Database searches  Database searches  Supplementary search  Supplementary search  Supplementary search  Supplementary search  Supplementary search  Database searches  Database searches  Database searches  Database searches  Database searches  Database searches  Database searches  Database searches  Supplementary search  Database searches  Database searches  Database searches  Supplementary search  Database searches  Database searches  Database searches  Database searches  Database searches  Database searches  Database searches  Database searches  Database searches  Database searches  Database searches  Database searches  Database searches  Database searches  Database searches  Supplementary search  Supplementary search  Database searches  Database searches  Database searches  Database searches  Database searches  Database searches  Database searches  Database searches  Database searches  Database searches  Database searches  Supplementary search  Database searches  Database searches  Database searches  Database searches  Database searches  Database searches  Database searches  Database searches | Outcome of interest  Publication type  Publication type  Publication type  Study design - Systematic review  Outcome of interest  Overlapping sample  Population type - homelessness status not clear  Population type  Study design  Outcome of interest  Population type - not homeless  Study design  Study design - Systematic review  Study design - No comparator  Study design - Systematic review  Study design - No comparator  Outcome of interest  Publication type  Overlapping sample  Overlapping sample  Publication type  Publication type – correction  Study design  Study design  Outcome of interest  Study design  Population type  Population type  Outcome of interest  Study design - Systematic review  Study design  Study design  Study design - Homelessness as covariate  Study design - Systematic review  Publication type – correction  Study design  Outcome of interest  Study design  Study design  Study design  Study design  Study design  Study design  Study design  Publication type  Study design  Population type  Study design  Outcome of interest  Publication type  Outcome of interest  Outcome of interest  Publication type  Outcome of interest  Study design - Systematic review  Insufficient data  Study design  Publication type  Outcome of interest  Outcome of interest  Outcome of interest  Study design - Cross sectional design  Study design  Publication type  Study design - Systematic review  Outcome of interest  Study design - No comparator  Study design - Systematic review  Population type  Study design  Study design  Publication type  Study design - Systematic review  Study design - Homelessness as covariate  Study design  Study design - Homelessness as covariate  Population type  Study design - Homelessness as covariate  Study design  Study design - Systematic review  Outcome of interest  Outcome of interest  Outcome of interest  Study design  Study design  Population type  Outcome  Study design - no comparator  Study design  Insufficient data  Study design - Systematic review  Study design  Study design - Homelessness as covariate  Population type  Insufficient data  Outcome of interest  Study design - Scoping review  Study design  Study design - No comparator  Publication type – correction  Publication type  Study design  Study design  Publication type  Study design - Systematic review  Study design  Publication type  Population type - not homeless  Population type  Outcome of interest  Study design  Population type  Study design  Outcome of interest  Publication type  Publication type  Insufficient data  Outcome of interest  Study design  Study design - No comparator  Outcome of interest  Study design  Study design  Study design - Homelessness as covariate  Insufficient data  Population type  Outcome of interest  Insufficient data  Publication type  Outcome of interest  Study design  Study design  Insufficient data  Outcome of interest  Outcome of interest  Population type - At 'risk' of homelessness  Study design  Publication type  Population type - not homeless  Study design - Cross sectional design  Study design  Outcome of interest  Study design  Outcome of interest  Publication type  Overlapping sample  Study design - Systematic review  Overlapping sample  Study design - Homelessness as covariate  Publication type  Study design  Study design  Outcome of interest  Study design - Homelessness as covariate  Outcome of interest  Study design  Outcome of interest  Study design  Publication type  Outcome of interest  Population type  Insufficient data  Population type  Outcome of interest  Study design  Study design  Outcome of interest  Study design  Study design  Publication type  Outcome of interest  Population type  Publication type  Population type  Study design - No comparator  Overlapping sample  Study design  Publication type  Outcome of interest  Outcome of interest  Insufficient data  Insufficient data  Population type - Homelessness status not clear  Study design - Research letters  Insufficient data  Publication type  Outcome of interest  Outcome of interest  Population type  Publication type  Publication type  Overlapping sample  Publication type  Study design  Publication type  Study design  Study design  Study design  Publication type  Study design  Overlapping sample  Study design  Study design  Insufficient data  Population type - Homelessness status not clear  Overlapping sample  Study design  Study design  Study design  Population type  Outcome of interest  Outcome of interest  Study design – No comparator  Study design  Study design  Outcome of interest  Study design  Study design - Systematic review  Study design  Population type - At 'risk' of homelessness  Study design  Study design  Publication type  Publication type  Population type – At ‘risk’ of homelessness  Outcome of interest  Study design  Publication type  Insufficient data  Study design  Study design  Population type  Population type |

### **Web appendix 5: Study characteristics**

*eTable 1. Characteristics of participants from published and unpublished studies*

| **Authors** | **Year** | **Number of participants** | | **N (%) homeless** | | **N (%) deaths in homeless** | **N (%) deaths in comparator** | | **Country** | **MASTER** | **Homelessness type** | **N (%) female** | **Mean age at baseline (years)** | **All-cause/**  **cause-specific mortality** |
| --- | --- | --- | --- | --- | --- | --- | --- | --- | --- | --- | --- | --- | --- | --- |
| **Published studies** |  | |  | |  |  | | |  |  |  |  |  |  |
| Agarwal ^12^ | 2019 | 8,575 | | 470 (5.5) | | 57 (12) | 496 (6.1) | | USA | 13 | NR | 3,517 (NR) | H: 45-64  C: 25-44 (mode) | All-cause |
| Alvarez-Uria ^13^ | 2013 | 4,105 | | 370 (9.0) | | - | - | | India | 13 | NR | 1,635 (40) | 25-34 (mode) | All-cause |
| Alvarez-Uria ^14^ | 2012 | 1,000 | | 46 (4.6) | | - | - | | India | 13 | NR | 341 (34) | 35 (median) | All-cause |
| Alvarez-Uria ^15^ | 2013 | 3,159 | | - | | - | - | | India | 12 | NR | 1,283 (41) | 25-35 (mode) | All-cause |
| Asaithambi ^16^ | 2021 | 364,922 | | 514 (0.1) | | 19 (3.7) | 31,714 (8.7) | | USA | 14 | NR | 181,247 (50) | H: 54.7  C:68.6 | All-cause |
| Averbuch ^17^ | 2022 | 4,287,478 | | 26,195 (0.6) | | 385 (1.5) | 218,580 (5.1) | | USA | 13 | NR | 2,088,219 (49) | 73.4 (median) | All-cause |
| Baggett ^18^ | 2013 | NR | | 28,033 (0.5) | | 1,278 (4.6) | NR | | USA | 15 | NR | NR | NR | Both |
| Balla^19^ | 2020 | 1,100,241 | | 3,938 (0.4) | | 240 (6.1) | 95,378 (8.7) | | USA | 13 | NR | 444,692 (40) | H: 17.5; C: 68 | All-cause |
| Barrow ^20^ | 1999 | NR | | 1,260 (0.0005) | | 161 (12.8) | NR | | USA | 10 | Shelter users | H: 311 (25) | 18-34 (mode) | All-cause |
| Baussano^21^ | 2008 | 1,564 | | 60 (3.8) | | 2 (3.3) | 104 (6.9) | | Italy | 12 | NR | 600 (38) | NR | NR |
| Beijer ^22^ | 2011 | NR | | 2,283 (0.1) | | 421 (18) | NR | | Sweden | 10 | NR | H: 526 (23) | 42.2 | Both |
| Bekele^23^ | 2018 | 454 | | 188 (41.4) | | 30 (16) | 23 (8.6) | | Canada | 12 | NR | 148 (NR) | 40-49 (mode) | All-cause |
| Beydoun ^24^ | 2024 | 6,128,921 | | 399,125 (6.5) | | NR | NR | | USA | 13 | NR | 475,624 (8) | 60-69 (mode) | Cause-specific |
| Bigé^25^ | 2015 | 9,774 | | 421 (4.3) | | 87 (20.7) | 2,251 (24.1) | | France | 10 | NR | H: 46 (11)  C: 3,992 (43) | H: 49; C: 62 (median) | All-cause |
| Borgdorff ^26^ | 1998 | 4,340 | | 55 (1.3) | | 4 (7.3) | 254 (5.9) | | Netherlands | 11 | NR | 1,737 (40) | 25-44 (mode) | Cause-specific |
| Braitstein ^27^ | 2021 | 1,321 | | 91 (6.9) | | 9 (9.9) | 3 (0.2) | | Kenya | 11 | Rough sleeping | 1,230 (NR) | 10.4 | All-cause |
| Brown ^28^ | 2022 | NR | | 450 (0.1) | | 117 (26) | NR | | USA | 11 | NR | H: 107 (24) | 58.1 (median) | All-cause |
| Capelastegui^29^ | 2023 | 16,172,792 | | 1,835 (0) | | 25 (1.4) | 141,028 (0.9) | | UK | 12 | NR | 8,654,323 (54) | 35.1 (median) | All-cause |
| Caylà ^30^ | 2004 | 1,217 | | 32 (2.6) | | 3 (9.4) | 20 (1.7) | | Spain | 13 | NR | 466 (NR) | 18-64 (mode) | All-cause |
| Chandra ^31^ | 2022 | 32,413 | | 150 (0.5) | | 12 (8) | 436 (1.4) | | USA | 12 | NR | 365 (NR) | Survivors: 65; Deceased: 68 (median) | All-cause |
| Chang ^32^ | 2022 | 7,610,551 | | 56,215 (0.7) | | 196 (0.3) | 217,219 (2.9) | | USA | 11 | NR | H: 2,270 (NR)  C: 4,890,980 (65) | 18-29 (mode) | Cause-specific |
| Chang ^33^ | 2023 | 16,859,189 | | 58,355 (0.3) | | 974 (1.7) | 88,658 (0.5) | | USA | 11 | NR | NR | H: 51-60 (mode) | All-cause |
| Chen ^34^ | 2018 | 161,124 | | 365 (0.2) | | 15 (4.1) | 5,646 (3.5) | | USA | 11 | NR | 50,239 (31) | 32.4 | All-cause |
| Clements ^35^ | 2022 | 52,903 | | 2,781 (5.3) | | 481 (17.3) | 5,595 (11.2) | | UK | 13 | NR | H; 975 (35) C: 28,973 (57.8) | 16NR24 | Both |
| Concannon ^36^ | 2020 | 81 | | 13 (16) | | 9 (69.2) | 51 (75) | | USA | 14 | Shelter users | H: 1 (7) C: 10 (15) | H:  Stage III/IV: 59.67,  C: Stage III/IV: 62.68 | Cause-specific |
| Courtepatte ^37^ | 2023 | 2,330 | | 415 (17.8) | | 18 (4.3) | 37 (1.9) | | USA | 17 | NR | 356 (15) | 26 (median) | All-cause |
| Dauby ^38^ | 2019 | 58 | | 28 (48.3) | | 3 (10.7) | 1 (3.3) | | Belgium | 10 | NR | 8 (14) | 43.7 | All-cause |
| Davidsen ^39^ | 2013 | NR | | 302 (0.01) | | 61 (20.2) | 112,935 (2.7) | | Denmark | 6 | NR | NR | NR | All-cause |
| Decker ^40^ | 2024 | 109,485 | | 5,356 (4.9) | | NR | NR | | USA | 15 | NR | H: 495 (9); C: 8,453 (8) | H: 63.1; C: 68.7 | All-cause |
| Decker ^41^ | 2023 | 111,267 | | 998 (0.9) | | 91 (9.1) | 7,917 (7.2) | | USA | 15 | NR | H: 288 (29)  C: 53,269 (48) | H: 49.9; C: 58 | All-cause |
| Demakakos ^42^ | 2020 | 6,366 | | 104 (1.6) | | 22 (21.2) | 1,064 (17) | | UK | 12 | NR | H: 57 (NR) C: 3,604 (NR) | H: 63.3; C: 60.7 | All-cause |
| Dibben ^43^ | 2011 | 13,303 | | 246 (1.8) | | 13 (5.3) | 326 (2.5) | | UK | 17 | NR | 4,050 (30) | 22.2 | All-cause |
| Feodor Nilsson^44^ | 2018 | 5,100,000 | | 29,058 (0.6) | | 4,345 (15) | 656,448 (12.9) | | Denmark | 13 | Shelter users | NR | NR | All-cause |
| Fine ^45^ | 2023 | NR | | 60,092 (0.1) | | 7,130 (11.9) | NR | | USA | 14 | NR | H: 22,008 (37) | 40.4 | Cause-specific |
| Fine ^46^ | 2020 | 5,948 | | 492 (8.3) | | 92 (18.7) | 572 (10.5) | | USA | 11 | NR | NR | 38.2 | Both |
| Gaber ^47^ | 2024 | 7,262 | | 637 (8.8) | | 51 (8) | 297 (4.5) | | Sweden | 18 | NR | H: 637 (NR) C: 2,361 (36) | H: 40.7  C: 47.1 | All-cause |
| Gromov ^48^ | 2022 | 436 | | 15 (3.4) | | 3 (20) | 12 (2.9) | | Russia | 11 | NR | NR | NR | Cause-specific |
| Haghighat^49^ | 2023 | 868 | | 151 (17.4) | | 27 (17.9) | 184 (25.7) | | USA | 10 | NR | H: 19 (13); C: 218 (30) | H: 56.7  C: 51.0 | Cause-specific |
| Hakansson ^50^ | 2013 | 4,081 | | NR | | NR | NR | | Sweden | 12 | NR | 407 (10) | Survivors: 33.4 Deceased 35.6 | All-cause |
| Haw ^51^ | 2006 | 10,825 | | 277 (2.6) | | 48 (17.3) | 1,128 (10.7) | | UK | 9 | NR | 6,639 (61) | 16-24 (mode) | Both |
| Henwood ^52^ | 2015 | 1,570,292 | | 292 (0.02) | | 41 (14) | NR | | USA | 12 | NR | H: 84 (29) | NR | All-cause |
| Hibbs ^53^ | 1994 | 1,408,049 | | 6,472 (0.5) | | 67 (1) | 6,568 (0.5) | | USA | 14 | NR | 3,795 (NR) | 51.3 | All-cause |
| Honer ^54^ | 2017 | NR | | 375 (0.001) | | 40 (10.7) | NR | | Canada | 14 | NR | 82 (22) | 34 (median) | All-cause |
| Hwang ^55^ | 2000 | NR | | 9,039 (0.8) | | NR | NR | | Canada | 13 | NR | 0 (0) | 43.7 | Both |
| Hwang ^56^ | 2009 | NR | | 15,100 (0.1) | | 3,280 (21.7) | NR | | Canada | 12 | Shelter users | 4,600 (30) | 36.1 | Both |
| Hwang ^57^ | 1997 | NR | | NR | | NR | NR | | USA | 8 | NR | 5,523 (50) | 25-44 (mode) | Both |
| Ivers ^58^ | 2019 | NR | | NR | | NR | NR | | Ireland | 12 | NR | 45 (22) | 42 (age at death) | All-cause |
| Jones ^59^ | 2015 | NR | | 371 (0.001) | | 31 (8.4) | NR | | Canada | 13 | NR | H: 81 (22) | 44 (median) | Both |
| Kasprow ^60^ | 2000 | 10,144 | | 6,714 (66.2) | | 1,045 (15.6) | 332 (9.7) | | USA | 13 | NR | 0 (0) | 43.9 NR 50.6 | All-cause |
| Kerker ^61^ | 2011 | NR | | 102,771 (1.3) | | 178 (0.2) | 175,505 (2.2) | | USA | 11 | NR | N not reported (69) | Gen. pop 12-17; Low-income 12-17;  Homeless 8-24 (mode) | All-cause |
| Khan ^61^ | 2021 | 1,900 | | 950 (50.0) | | 63 (6.6) | 45 (4.7) | | USA | 16 | NR | H: N (35) C: N (41) | Homeless: 41, Not-homeless: 61 | All-cause |
| Khan ^62^ | 2023 | 648,944 | | 3,134 (0.5) | | 282 (9) | 63,046 (9.8) | | USA | 11 | NR | H: N (33) C: N (49) | Homeless: 39, Not-homeless: 38 (median) | All-cause |
| Khanijow ^63^ | 2015 | 5,472 | | 558 (10.2) | | NR | NR | | USA | 11 | NR | 408 (7) | Mode: 30-39 | All-cause |
| Kiwanuka ^64^ | 2019 | 43,872 | | 332 (0.8) | | 11 (3.3) | 1,785 (4.1) | | USA | 14 | NR | H: 39 (12) C: 13,244 (30) | Homeless: 46.1, Non-homeless: 46.0 | All-cause |
| Koyama ^65^ | 2024 | 836,361 | | 46,561 (5.6) | | NR | NR | | USA | 18 | NR | H: 2,326 (5) C: 24,451 (3) | Homelessness: 55-64;  Not homeless 65-74 (mode) | All-cause |
| Kurbatova ^66^ | 2012 | 1,174 | | 18 (1.5) | | 2 (11.1) | 198 (17.1) | | Estonia, Latvia, Philippines, Russia, Peru | 8 | NR | 493 (NR) | 36 (median) | Cause-specific |
| Lebow ^67^ | 1995 | 1,608 | | 72 (4.5) | | 30 (41.7) | 553 (36) | | USA | 10 | NR | H: 13 (18)  C: 166 (11) | 30-39 (mode) | All-cause |
| Lee ^68^ | 2019 | 168 | | 56 (33.3) | | 13 (23.2) | 23 (20.5) | | Korea | 12 | NR | H: 3 (5) C: 6 (5) | H: 54.4  C: 54.0 | All-cause |
| Lemay ^69^ | 2019 | 1,689 | | 319 (18.9) | | 22 (6.9) | 185 (13.5) | | Canada | 13 | NR | NR | H: 45  C: 57.9 | All-cause |
| Levanon Seligson^70^ | 2017 | 244,298 | | 27,441 (11.2) | | NR | NR | | USA | 10 | Shelter users | Incarcerated: N (10)  Comparator 1: N (54)  Comparator 2: N (56) | Incarcerated: 35-49 (mode) | All-cause |
| Liauw ^71^ | 2021 | 2,854 | | 75 (2.6) | | 14 (18.7) | 155 (5.6) | | Canada | 14 | NR | H: 3 (4); C: 671 (24) | H: 58  C: 63 | All-cause |
| Lim ^72^ | 2012 | 155,272 | | 16,216 (10.4) | | NR | NR | | USA | 12 | Shelter users | 18,111 (12) | 16-24 (mode) | Cause-specific |
| Lopes ^73^ | 2024 | 6,581 | | 189 (2.9) | | 18 (9.5) | 263 (4.1) | | Brazil | 15 | NR | 3,895 (NR) | 18-39 (mode) | Cause-specific |
| Miller ^74^ | 2020 | 393 | | 131 (33.3) | | 0 (0) | 6 (2.3) | | Australia | 12 | NR | H: 15 (11)  C: 30 (11) | H: 53.4  C: 53.5 | All-cause |
| Miller-Archie ^75^ | 2022 | 2,173 | | 1,312 (60.4) | | 232 (17.7) | 153 (17.8) | | USA | 15 | NR | NR | H: 45-64,  C: 45-64 (mode) | Both |
| Miyawaki ^76^ | 2020 | 20,078 | | 1,295 (6.4) | | 18 (1.4) | 465 (2.5) | | USA | 12 | NR | NR | 40 (median) | Both |
| Munteanu ^77^ | 2022 | 1,123 | | 26 (2.3) | | 7 (26.9) | 192 (17.5) | | Romania | 9 | NR | 12,938 (NR) | 40-59 (mode) | All-cause |
| Nanjo ^78^ | 2020 | 40,626 | | 8,492 (20.9) | | NR | NR | | UK | 11 | NR | H: 3,517 (41) C: 13,484 (42) | H: 39.0  C: 38.3 | Cause-specific |
| Nathanson ^78^ | 2019 | 781,540 | | 2,278 (0.3) | | 109 (4.8) | 65,458 (8.4) | | USA | 13 | NR | H: 4408 (19) C: 363,915 (47) | H: 47.8 | Cause-specific |
| Nicholas ^79^ | 2021 | NR | | 166,749 (1.6) | | 3,376 (2) | NR | | USA | 11 | NR | NR | NR | Both |
| Nicholas ^80^ | 2021 | 22,459,252 | | 810,326 (3.6) | | 104,418 (12.9) | 2,453,430 (11.3) | | USA | 6 | NR | H: 259,991 (32) C:10,143,568 (47) | H: 18-64  C: 18-64 (mode) | All-cause |
| Nilsson ^81^ | 2022 | 249,924 | | 269 (0.1) | | 6 (2.2) | 2,857 (1.1) | | Denmark | 15 | Shelter users | 127,335 (51) | 40 (median) | All-cause |
| Nordentoft ^82^ | 2003 | NR | | 559 (0.1) | | 141 (25.2) | NR | | Denmark | 10 | Shelter users | NR | NR | Both |
| Nusselder ^83^ | 2013 | NR | | 2,096 (0.3) | | 266 (12.7) | NR | | Netherlands | 11 | NR | H: 250 (12) | 40.6 | All-cause |
| O'Driscoll ^84^ | 2001 | 2,849 | | 1,009 (35.4) | | 33 (3.3) | 35 (1.2) | | USA | 14 | NR | 1,051 (37) | 36.5 | Both |
| Paniagua-Saldarriaga^85^ | 2021 | 5,698 | | 269 (4.7) | | 28 (10.4) | 519 (9.6) | | Colombia | 14 | NR | (46) | 25-34 (mode) | All-cause |
| Park ^86^ | 2024 | 8,450 | | 632 (7.5) | | 31 (4.9) | 464 (5.9) | | USA | 18 | NR | 3,089 (20) | N/S | All-cause |
| Peak ^87^ | 2020 | 835 | | 544 (65.1) | | 253 (46.5) | 14 (4.8) | | USA | 11 | NR | 189 (32) | 43 | Cause-specific |
| Pradipta ^88^ | 2019 | 5,674 | | 132 (2.3) | | 5 (3.8) | 117 (2.1) | | Netherlands | 13 | NR | 2,116 (38) | 25-74 (mode) | Cause-specific |
| Pradipta ^89^ | 2019 | 545 | | 15 (2.8) | | 0 (0) | 6 (1.1) | | Netherlands | 13 | NR | 250 (46) | 25-64 (mode) | Cause-specific |
| Ranzani ^90^ | 2020 | 15,342 | | 376 (2.5) | | 119 (31.6) | 2,382 (15.9) | | Brazil | 15 | NR | NR | 25-35 (mode) | Both |
| Ranzani ^91^ | 2016 | 61,817 | | 1,726 (2.8) | | 181 (10.5) | 3,619 (6) | | Brazil | 15 | NR | 17,245 (NR) | 25-35 (mode) | All-cause |
| Rayburn ^91^ | 2012 | NR | | 670 (0.1) | | 94 (14) | 83 (0) | | USA | 11 | NR | H 167 (25) | 32.4 | All-cause |
| Richard ^92^ | 2024 | 7,040 | | 640 (9.1) | | 17 (2.7) | 46 (0.7) | | Canada | 18 | Shelter users | NR | - | All-cause |
| Richard ^93^ | 2021 | 28,704 | | 274 (1.0) | | 10 (3.6) | 730 (2.6) | | Canada | 9 | NR | 7,833,506 (NR) | H: 38  C: 41 (median) | Cause-specific |
| Roncarati ^94^ | 2018 | 4,849,478 | | 445 (0.009) | | 134 (30.1) | NR | | USA | 11 | Rough sleeper | 25,568,906 (NR) | 18-44 (mode) | Both |
| Rousssos ^95^ | 2024 | 2,428 | | 463 (19.1) | | 66 (14.3) | 176 (9) | | Greece | 10 | NR | 375 (15) | 40.1 | All-cause |
| Roy ^96^ | 2010 | NR | | 2,694 (0) | | 57 (2.1) | NR | | Canada | 10 | NR | NR | Cohort 1: 20.4 yrs Cohort 2: 19.9 yrs | All-cause |
| Sadowski ^97^ | 2009 | 407 | | 206 (50.6) | | 23 (11.2) | 25 (12.4) | | USA | 32 | Shelter users | 95 (23) | Intervention: 47 Usual Care: 46 | All-cause |
| Saitz ^98^ | 2007 | 470 | | 221 (47) | | NR | NR | | USA | 16 | NR | 113 (24) | 35 (median) | All-cause |
| Schinka ^99^ | 2018 | 89,096 | | 23,898 (26.8) | | 3,905 (16.3) | 4,143 (6.4) | | USA | 12 | NR | H: 3.4, C: N 3.6 | H: 45.1 | Both |
| Schinka ^100^ | 2016 | 24,546 | | 4,475 (18.2) | | 1,560 (34.9) | 3,649 (18.2) | | USA | 14 | NR | H: 49 (1); C: 604 (3) | H: 59.2  C: 59.4 | All-cause |
| Schmit ^101^ | 2020 | 2,273 | | 325 (14.3) | | 35 (10.8) | 160 (8.2) | | USA | 10 | NR | 660 (29) | 45-64 (mode) | All-cause |
| Schwarcz ^102^ | 2009 | 6,569 | | 652 (9.9) | | 211 (32.4) | 431 (7.3) | | USA | 15 | NR | 493 (8) | 30-39 (mode) | Both |
| Scott ^103^ | 2023 | NR | | NR | | 1,271 (N) | NR | | USA | 10 | NR | 235 (18) | 45-64 (mode) | Cause-specific |
| Seastres ^104^ | 2020 | 6,300 | | 1,575 (25) | | 237 (15) | 498 (10.5) | | Australia | 15 | NR | 2,583 (41) | H: 41.02  C: 40.27 (median) | All-cause |
| Shaw ^105^ | 1999 | NR | | 1,866 (0.01) | | 210 (11.3) | NR | | UK | 6 | Rough sleepers  Hostel residents Bed and breakfast/ bedsit residents | Rough sleepers Hostel residents 0 (all male)  Bed and breakfast/ bedsit residents: 297 (32) | Rough sleepers: 16-29  Hostel residents: 16-44  B&B: 16-44 | All-cause |
| Skicki ^106^ | 2022 | 355,097 | | 773 (0.2) | | 103 (13.3) | 21,295 (6) | | USA | 15 | NR | H: 136 (NR) C: 144,210 (NR) | H: 47.0, C: 54.0 | All-cause |
| Slockers ^107^ | 2018 | NR | | 2,126 (0.3) | | 261 (12.3) | NR | | Netherlands | 15 | NR | H: 260 (12) | 40.3 | Cause-specific |
| Smith ^108^ | 2017 | 126 | | 63 (50) | | 18 (28.6) | 5 (7.9) | | Canada | 10 | NR | 63 (NR) | 48 | All-cause |
| Stenius-Ayoade^109^ | 2017 | 1,857 | | 617 (33.2) | | 287 (46.5) | 138 (11.1) | | Finland | 14 | Shelter users | 0 (0) | 49 | Both |
| Subramanian ^110^ | 2022 | 16,332 | | 4,083 (25) | | 137 (3.4) | 302 (2.5) | | USA | 10 | NR | Upper GI: H:31  C: 43 Lower GI: H: 33; C: 32 | Upper GI:  H: 57.7  C: 66.8 Lower GI:  H: 65.2  C: 72.5 years | All-cause |
| Tsai ^111^ | 2024 | 6,128,921 | | 399,125 (6.5) | | 3,897 (1) | 5,427 (0.1) | | USA | 18 | NR | NR | - | Cause-specific |
| Tweed ^112^ | 2022 | 518,004 | | 9,463 (1.7) | | 241 (2.5) | 10,103 (1.9) | | UK | 14 | NR | 47 | 40.5 | All-cause |
| van Laere ^113^ | 2009 | NR | | 517 (0.1) | | 83 (16.1) | NR | | Netherlands | 6 | Shelter users | 109 (NR) | 40-49 (mode) | Both |
| Vila-Rodriguez ^114^ | 2013 | NR | | 293 (0.001) | | 15 (5.1) | NR | | Canada | 11 | NR | 68 (23) | 44.1 (median) | All-cause |
| Wadhera ^115^ | 2019 | 32,507,861 | | 185,292 (0.6) | | 1,668 (0.9) | 381,406 (1.2) | | USA | 12 | NR | H: 24; C: 24 | H: 46.1  C: 46.1 | All-cause |
| Walley ^116^ | 2008 | 595 | | 164 (27.6) | | NR | NR | | USA | 11 | NR | 148 (25) | 41 | All-cause |
| Wang ^117^ | 2020 | 3,740 | | 372 (9.9) | | 3 (0.8) | 918 (27.3) | | Canada | 11 | Shelter users | 9,150 (NR) | H: <50  C: 80+ (mode) | Cause-specific |
| White ^118^ | 2022 | 23,678 | | 1,453 (6.1) | | 70 (4.8) | 735 (3.3) | | UK | 12 | Rough, Shelter users, squat, sofa surf | 11,997 (51) | 1958 cohort: 23-58  1970 cohort: 16-44 (mode) | All-cause |
| Zagdyn ^119^ | 2017 | 2,888 | | 137 (4.7) | | 46 (33.6) | 609 (22.1) | | Russia | 12 | NR | 712 (25) | 32.9 | Cause-specific |
| Zagozdzon ^120^ | 2016 | 47,247 | | 816 (1.7) | | 23 (2.8) | 957 (2.1) | | Poland | 10 | NR | H: 248 (30) C: 22,890 (49) | H: 45.9  C: 45.5 | Both |
| Zivanovic ^121^ | 2015 | 2,435 | | 1,713 (70.3) | | NR | NR | | Canada | 12 | NR | 833 (34) | 37.7 (median) | All-cause |
| Zordan ^122^ | 2023 | 5,772 | | 1,050 (18.2) | | 382 (36.4) | 1,204 (25.5) | | Australia | 14 | NR | 2,581 (NR) | H: 38; Housed: 40; Marginally housed:49 (median) | All-cause |
| **Unpublished studies** |  | |  | |  |  | | |  |  |  |  |  |  |
| HRS | 2024 | 7,250 | | 382 (5.3) | | 73 (19.1) | 1,632 (23.8) | | USA | 15 | Shelter users | 3,939 (NR) | 67 | All-cause |
| SHARE | 2024 | 24,233 | | 83 (0.3) | | 20 (24.1) | 4,841 (20) | | Europe | 15 | NR | 13,606 (56) | 66.1 | All-cause |
| **Total** |  | **131,464,809** | | **2,563,633 (2.0%)** | | **146,147 (5.7%)** | |  |  |  |  |  |  |  |
| **All-cause** |  | **110,563,112** | | **1,618,049 (1.5%)** | | **132,903 (8.2%)** | |  |  |  |  |  |  |  |
| **Cause-specific** |  | **26,006,062** | | **1,202,205 (0.5%)** | | **27,437 (2.3%)** | |  |  |  |  |  |  |  |

NR: Not reported; H: Homeless; C: Comparator

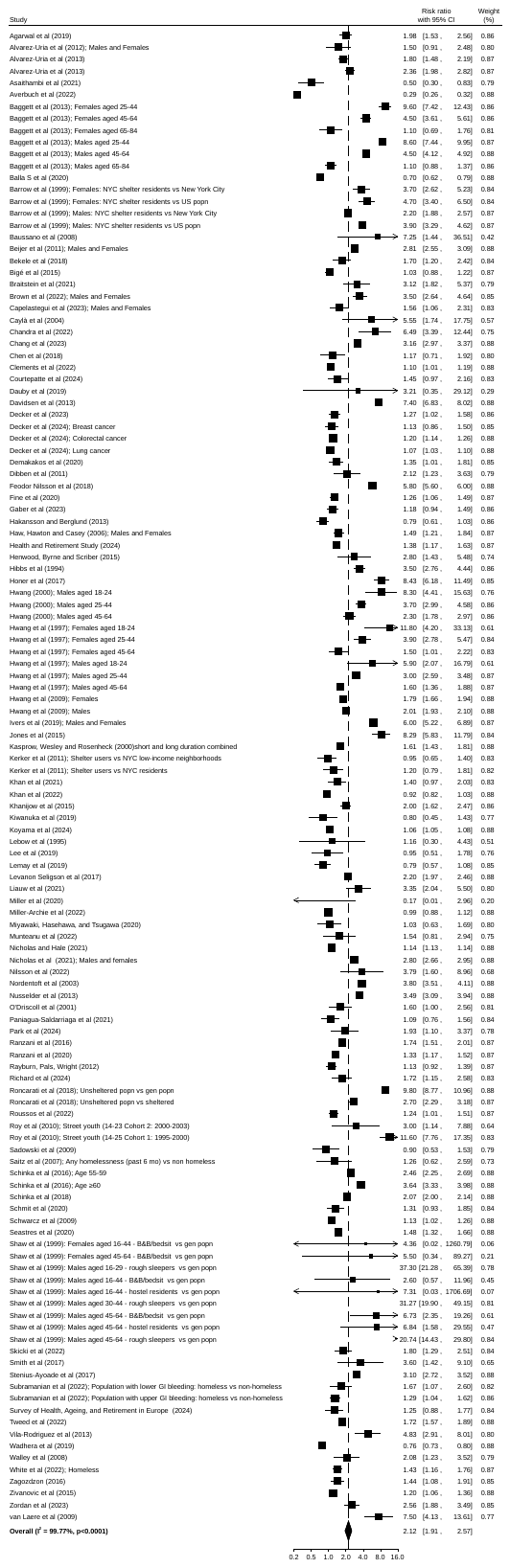

### **Web appendix 6**

**Figure 1: Forest plot of RRs for all-cause mortality**

Several studies contributed multiple effect sizes because different exposed groups, comparator groups or length follow-ups were reported. 126 independent effect estimates were reported in 95 studies. Any correlation between RRs has been accounted for in the summary RR. The most comprehensively adjusted estimates were chosen for all studies. RR= relative risk.

0

1

2

3

-4

-2

0

2

4

6

Effect size

95% CI

Summary relative risk

Study relative risks

Standard error

### **Web appendix 6: Figure 2. Funnel plot for association of homelessness with all-cause mortality**

### **Web appendix 8: MOOSE (Meta-analyses Of Observational Studies in Epidemiology) checklist**

| **Item No** |  | **Reported on Page No** |
| --- | --- | --- |
| **Reporting of background should include** | | |
| 1 | Problem definition | 6 |
| 2 | Hypothesis statement | 6-7 |
| 3 | Description of study outcome(s) | 9 |
| 4 | Type of exposure or intervention used | 8 |
| 5 | Type of study designs used | 8 |
| 6 | Study population | 7-8 |
| **Reporting of search strategy should include** | | |
| 7 | Qualifications of searchers (eg, librarians and investigators) | 7 |
| 8 | Search strategy, including time period included in the synthesis and key words | Appendix |
| 9 | Effort to include all available studies, including contact with authors | 7 |
| 10 | Databases and registries searched | 7, Appendix |
| 11 | Search software used, name and version, including special features used (eg, explosion) | 11 |
| 12 | Use of hand searching (eg, reference lists of obtained articles) | 7-8 |
| 13 | List of citations located and those excluded, including justification | Appendix |
| 14 | Method of addressing articles published in languages other than English | 7 |
| 15 | Method of handling abstracts and unpublished studies | 7 |
| 16 | Description of any contact with authors | 8 |
| **Reporting of methods should include** | | |
| 17 | Description of relevance or appropriateness of studies assembled for assessing the hypothesis to be tested | 7-8 |
| 18 | Rationale for the selection and coding of data (eg, sound clinical principles or convenience) | 7-8 |
| 19 | Documentation of how data were classified and coded (eg, multiple raters, blinding and interrater reliability) | 7-8 |
| 20 | Assessment of confounding (eg, comparability of cases and controls in studies where appropriate) | 8 |
| 21 | Assessment of study quality, including blinding of quality assessors, stratification or regression on possible predictors of study results | 9 |
| 22 | Assessment of heterogeneity | 10 |
| 23 | Description of statistical methods (eg, complete description of fixed or random effects models, justification of whether the chosen models account for predictors of study results, dose-response models, or cumulative meta-analysis) in sufficient detail to be replicated | 10 |
| 24 | Provision of appropriate tables and graphics | Figures 1-4, Appendix |
| **Reporting of results should include** | | |
| 25 | Graphic summarizing individual study estimates and overall estimate | eFigure 1 Appendix |
| 26 | Table giving descriptive information for each study included | Appendix |
| 27 | Results of sensitivity testing (eg, subgroup analysis) | 14 |
| 28 | Indication of statistical uncertainty of findings | 13-14 |
